# Prevalence and Characteristics of New Mental Health Interventions in PICU Survivors

**DOI:** 10.1101/2025.09.20.25336236

**Authors:** Mariah L DeSerisy, Julia A. Heneghan, Matt Hall, Daniel H. Choi, Leslie A. Dervan, Daniel Garros, Denise M. Goodman, Jason M. Kane, Joseph G. Kohne, Colin M. Rogerson, Nadia Roumeliotis, Vanessa Toomey, Adam C. Dziorny

## Abstract

**Background and Objectives:** As survival after pediatric critical illness improves, attention has shifted to post-intensive care syndrome (PICS-p) and specifically the long-term mental health of PICU survivors, who face elevated risks including posttraumatic stress, anxiety, and depression. However, little is known about actual patterns of post-discharge mental health care. The objective of this study is to examine the rates of mental health follow-up and psychopharmacology use among publicly insured children following PICU hospitalization, compared with those hospitalized on acute care wards, using a multi-state administrative dataset.

**Methods:** We performed a retrospective cohort study using 2016–2021 Medicaid claims across 10–12 states. The cohort comprised children aged 3–18 years discharged home after an index hospitalization and excluded perinatal admissions and hospitalizations primarily for mental health or traumatic brain injury. The primary exposure was pediatric intensive care unit (PICU) admission. The primary outcome was new mental health visits within one-year post-discharge. Secondary outcomes included visit provider type, visit diagnoses category, and new psychiatric prescriptions. We report descriptive statistics and measure associations with covariates using logistic regression.

**Results:** Among 144,763 Medicaid-insured pediatric hospitalizations (20.7% with PICU stays), only 8.8% initiated new mental health care. When compared to hospitalizations without PICU exposure, those with PICU exposure were more likely to complete new mental health visits (n=1,697 [6.1%] of PICU hospitalizations vs 5,252 [4.9%] of non-PICU hospitalizations). However, PICU exposure was not independently associated with a new mental health visit after adjustment (OR 1.06, 95% CI 1 – 1.13; p=0.067). Older age, complex chronic conditions, and longer length of stay were associated with new mental health visits. Hospitalizations with a PICU stay were significantly associated with increased rate of visits to psychologists or supportive therapists compared to those without a PICU stay (p<0.001).

**Conclusions:** Mental health follow-up after pediatric hospitalization is rare. Future studies should investigate barriers to care and identify effective methods for systematic screening and proactive referral.

## Introduction

Childhood critical illness accounts for nearly 1 in 6 childhood hospitalizations.^1^ As more children survive critical illness,^2^ research has shifted to understand the mental and emotional impact of hospitalization on children and their families to improve post-hospitalization outcomes and quality of life.^3–5^ Post-intensive care syndrome in pediatrics (PICS-p) describes a framework for understanding common impairments following PICU hospitalization, taking into account children’s developmental status and the effects of hospitalization on both children and families.^3^ The framework proposes four interrelated core domains: physical health, emotional health, cognitive health, and social health.^3^

Meta-analyses describe significant, long lasting emotional burdens following PICU discharge,^6^ including high rates of acute posttraumatic stress, anxiety, and depression among survivors.^7–9^ One recent study leveraging a large commercial payor database found that, compared to children who had never been hospitalized, PICU survivors demonstrated a 63% increase in odds of a new mental health disorder diagnosis within one year post-discharge.^10^ If these mental health challenges remain untreated, they are associated with poor quality of life,^11,12^ lower functional capacity,^13,14^ and increased health care utilization.^13^ Previous work identifying the high prevalence of mental health symptoms and new diagnoses among PICU survivors suggests that screening and effective referral programs are necessary for PICU survivors. Despite this, no research has yet examined current rates and details of mental health care (e.g., medication, individual therapy provider types) received by PICU survivors and their families.

This study aims to examine associations between mental health follow-up and PICU survivorship, adjusting for patient features and clinical characteristics. We assessed the rate of mental health practitioner visits following PICU admission using a multi-state administrative dataset. This study uniquely focused on children with public (Medicaid) insurance, quantified specific follow-up visit utilization, and measured filled psychopharmacotherapy prescriptions, including antidepressants and anxiolytics. We chose this population due to our interest in this high-risk population and the availability of high-quality longitudinal claims data. We hypothesized that patients with hospitalization including a PICU stay would receive increased mental health follow-up, and more frequent filled psychopharmacology prescriptions, compared to patients hospitalized on an acute care ward. Better understanding of current rates of use and types of mental health treatment received by survivors and families may identify gaps in current practice, thereby informing more effective screening and referral practices post-PICU discharge to ultimately reduce psychiatric morbidity.

## Methods

This study followed the Strengthening the Reporting of Observational Studies in Epidemiology reporting guidelines for observational studies (Supplementary Table 1).^15^ This study was deemed not human subjects research by the University of Rochester Institutional Review Board (#STUDY00010686).

### Setting and design

We analyzed administrative claims data from the Marketscan Medicaid claims database (IBM Watson Health, Armonk, New York). The database includes standardized, integrated, enrollee-level, de-identified claims across inpatient and outpatient settings as well as prescription medication services for both fee-for-service and managed care plans from 10 to 12 geographically dispersed states, depending on the year. The database includes de-identified demographic data about the enrollees, their medical providers, and healthcare facilities. An encrypted person identifier allows for longitudinal follow-up of enrollees over time.

### Patient population

Our unit of analysis was an index hospitalization occurring between 2016 – 2021. We included children between the ages of 3 and 18 years who were discharged home from their index hospitalization. We included preschool children as young as 3 years of age consistent with prior epidemiologic studies of mental health care among children.^10,16^ We included a one-year “look-back” period to assess for mental health care in the year prior to the index hospitalization. We excluded patients born in the hospital or admitted from the NICU, as well as patients who did not have one year of pre-or post-enrollment in Medicaid surrounding the index hospitalization. We excluded primary hospitalizations within Major Diagnosis Category 14, referring to pregnancy, childbirth, and the perinatal period. Because we were interested in the rate of new mental health follow-up post-PICU discharge, we excluded patients with primary admission diagnosis of a mental health concern (e.g., suicide attempt) or of traumatic brain injury (TBI),^17^ as these conditions have existing pathways for mental health linkage.^18,19^ Only subjects with complete data (e.g., no missingness) were included. A patient flow diagram is provided in Supplementary Figure 1.

### Measurements

Our primary outcome was the rate of new mental health visits in the one year following the index hospitalization. We defined mental health visits as those billed by any primary mental health care provider, including psychiatric prescribers (e.g., psychiatrists, psychiatric nurse practitioners) and therapists (e.g., supportive therapists, licensed clinical social workers, psychologists) for a mental health diagnosis as defined by Child and Adolescent Mental Health Disorders Classification System diagnosis codes.^20^ Consistent with prior studies of mental health treatment post PICU care, mental health visits regarding the child but conducted with parents (i.e., parent training or family-focused care) were included.^21^ To evaluate new mental health visits, we excluded patients with mental health care visits in the year prior to the index hospitalization. Secondary outcomes included rates of psychiatric medications prescribed, as identified by manual review of the Marketscan prescription therapeutic class.

We defined the primary exposure as a PICU stay during the index hospitalization, defined by intensive care billing codes. We conducted a comparison of hospitalizations with and without a PICU stay on our primary outcome of MH visits. Covariates included the number of chronic complex conditions (CCCs) as defined by billing codes,^22^ index hospitalization length of stay in days, the need for mechanical ventilation and/or continuous renal replacement therapy (see codes in Supplementary Table 2),^23,24^ and the primary reason for hospital admission grouped by Pediatric Clinical Classification System (PECCS) code.^25^

### Analysis

We performed descriptive analysis of categorical data with frequencies and percentages; two sample t-tests were used to compare distributions of continuous variables between groups (i.e., PICU hospitalization versus general care hospitalization). Among those patients who had new mental health utilization following admission, we described the rate of mental health visits by provider type and the visit diagnoses. We also performed bivariate analysis of PICU admission status with new psychopharmaceutical prescriptions following hospital admission. We performed a primary multivariable logistic regression analysis measuring the association of the exposure (PICU admission) with the outcome of new post-hospitalization mental health visits, adjusted for patient and clinical covariates. Pre-planned secondary analyses measured associations to two additional secondary outcomes: new internalizing (e.g., anxiety, depression, and trauma-related) and new externalizing (e.g., ADHD, disruptive, and conduct) mental health diagnoses,^26,27^ coded from diagnoses codes applied to follow-up MH visits. We calculated odds ratios, with 95% CIs, using logistic regression models. All analyses were performed using SAS Version 9.4 (SAS Institute, Cary, NC). A p value of less than 0.05 was considered significant.

## Results

### Patient Characteristics

We identified 144,763 index hospitalizations that met inclusion criteria. Among these, 29,994 (20.7%) included a PICU stay. Overall, 8.8% (n=12,739) of all included children received mental health follow-up in the 12 months following the index hospitalization. After excluding patients with mental health utilization in the one year prior to admission, 5.2% (n=6,949) of patients received new mental health care. Primary admission diagnosis is indicated in Supplementary Table 3.

Patients with new mental health utilization post-hospitalization were equally likely to be male or female. All remaining demographic features, such as age, race, comorbid conditions, and length of stay were significantly associated with post-hospital mental health utilization (Table 1). New-mental health visit patients were more likely to be older, to have complex chronic conditions, to have a Medicaid basis of eligibility from being blind or disabled, and to have an increased hospital length of stay. Hospital length of stay demonstrated a dose-response relationship, with increasing proportion of new mental health visits for increased length of stay among both PICU-hospitalized and non-PICU hospitalized patients.

**Table 1.** Demographics of the subject cohort stratified by exposure to PICU hospitalization, and outcome of mental health visits.

|  |  | Overall<br>(N=144,763) | Non-PICU Hospitalizations<br>(N=114,769, 79.3%) |  | PICU Hospitalizations<br>(N=29,994, 20.7%) |  |
| --- | --- | --- | --- | --- | --- | --- |
|  |  |  | No Post-Hospital<br>MH Visits<br>N=104,950, 91.4% | Post-Hospital MH<br>Visits<br>N=9,819, 8.6% | No Post-Hospital<br>MH Visits<br>N=27,074, 90.3% | Post-Hospital MH<br>Visits<br>N=2,920, 9.7% |
| **Age (years)** | 3 – 5 | 40,025 (27.6) | 30,112 (28.7) | 1,623 (16.5) | 7,783 (28.7) | 507 (17.4) |
|  | 6 – 10 | 40,892 (28.2) | 30,079 (28.7) | 2,853 (29.1) | 7,197 (26.6) | 763 (26.1) |
|  | 11 – 14 | 30,246 (20.9) | 21,228 (20.2) | 2,563 (26.1) | 5,674 (21) | 781 (26.7) |
|  | 15 – 18 | 33,600 (23.2) | 23,531 (22.4) | 2,780 (28.3) | 6,420 (23.7) | 869 (29.8) |
| **Sex** | Female | 67,620 (46.7) | 49,702 (47.4) | 4,614 (47) | 12,050 (44.5) | 1,254 (42.9) |
| **Race** | White | 66,782 (46.1) | 49,385 (47.1) | 4,906 (50) | 11,146 (41.2) | 1,345 (46.1) |
|  | Black | 49,489 (34.2) | 35,024 (33.4) | 2,990 (30.5) | 10,501 (38.8) | 974 (33.4) |
|  | Hispanic | 8,965 (6.2) | 6,470 (6.2) | 638 (6.5) | 1,652 (6.1) | 205 (7) |
|  | Other | 4,865 (3.4) | 3,657 (3.5) | 301 (3.1) | 797 (2.9) | 110 (3.8) |
| **Pre-Hospitalization MH** | Yes | 10,297 (7.1) | 3,486 (3.3) | 4,567 (46.5) | 1,021 (3.8) | 1,223 (41.9) |
| **Number of CCCs** | 0 | 94,249 (65.1) | 72,821 (69.4) | 6,193 (63.1) | 13,870 (51.2) | 1,365 (46.7) |
|  | 1 | 34,734 (24) | 22,731 (21.7) | 2,609 (26.6) | 8,357 (30.9) | 1,037 (35.5) |
|  | 2 | 11,742 (8.1) | 7,191 (6.9) | 782 (8) | 3,396 (12.5) | 373 (12.8) |
|  | 3+ | 4,038 (2.8) | 2,207 (2.1) | 235 (2.4) | 1,451 (5.4) | 145 (5) |
| **Blind or Disabled <sup>a</sup>** | Yes | 17,898 (12.4) | 11,292 (10.8) | 1,931 (19.7) | 4,122 (15.2) | 553 (18.9) |
| **Hospital Length of Stay** | 0 – 2 days | 83,071 (57.4) | 64,982 (61.9) | 5,893 (60) | 11,003 (40.6) | 1,193 (40.9) |
|  | 3 – 7 days | 51,522 (35.6) | 35,062 (33.4) | 3,277 (33.4) | 11,997 (44.3) | 1,186 (40.6) |
|  | 8 – 14 days | 7,128 (4.9) | 3,801 (3.6) | 448 (4.6) | 2,597 (9.6) | 282 (9.7) |
|  | 15 – 30 days | 2,311 (1.6) | 930 (0.9) | 137 (1.4) | 1,071 (4) | 173 (5.9) |
|  | 31+ days | 731 (0.5) | 175 (0.2) | 64 (0.7) | 406 (1.5) | 86 (2.9) |

All percentages are given as percentage of the column sample except in the header rows, where they are percentage of the larger population.
All features are significantly associated with Post-Hospital MH visits, within each PICU vs Non-PICU Hospitalization ( $p < 0.001$ ), except for sex.
Abbreviations: PICU, pediatric intensive care unit; MH, mental health; CCC, complex chronic conditions
<sup>a</sup> Indicated by a Basis of Eligibility code equal to “Blind / Disabled Individual” as opposed to “Child” codes for eligibility, including “Child of Unemployed Adult” and “Foster Care Child”

### Primary Outcome

When compared to patients hospitalized without PICU exposure and excluding those with prior mental health care, patients with PICU exposure were more likely to participate in a new mental health care visit in the 12 months following the index hospitalization (n=1,697 [6.1%] of PICU hospitalizations vs 5,252 [4.9%] of non-PICU hospitalizations, Table 2). Similar differences were observed when we included those with prior mental health care (Supplementary Table 4).

**Table 2.**
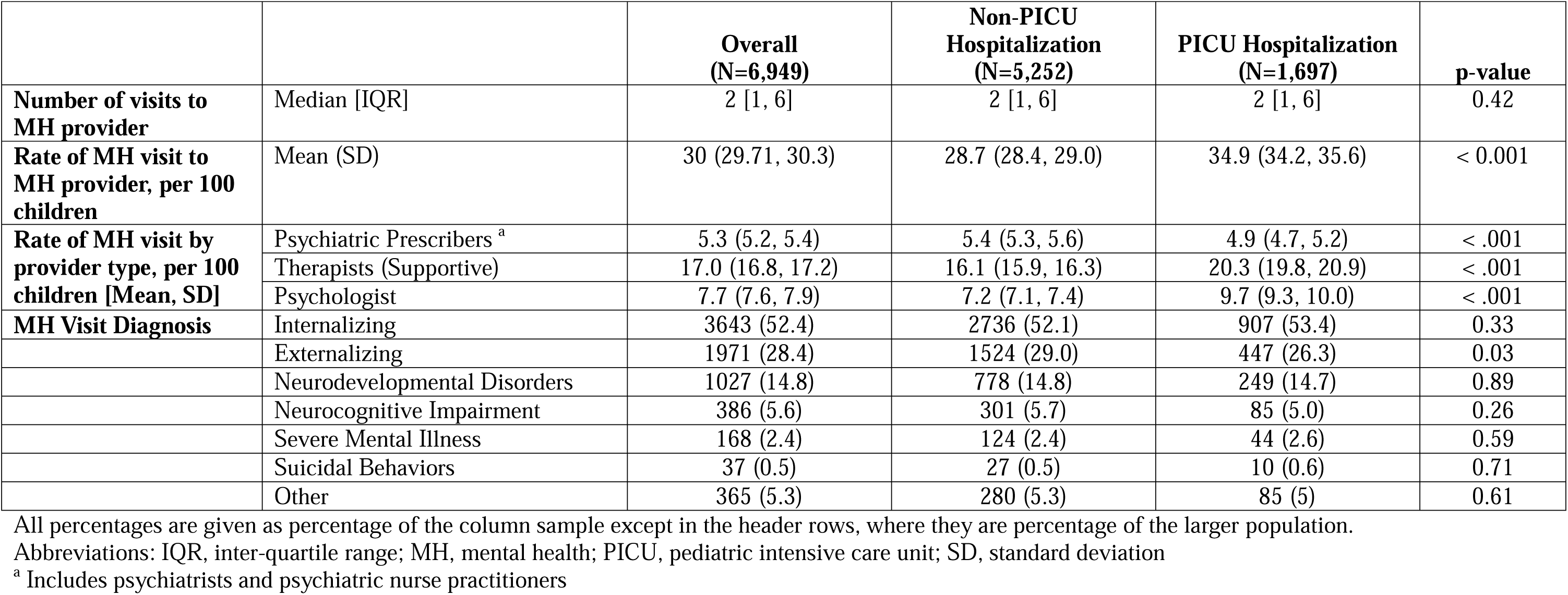
New mental health provider utilization in the one year following index admission, excluding patients with MH visits in the 1-year prior to index hospitalization.

Age, complex chronic conditions, and hospital length of stay were independently associated with new mental health utilization in a multivariable regression model adjusting for PICU exposure, sex, number of CCCs, and level of support received (Table 3). PICU exposure itself was not independently associated with new mental health care in this model (OR 1.06 [95% CI: 1.0 – 1.13], p=0.067, Table 3).

**Table 3.** Multivariable logistic regression model of 12-month new mental health utilization outcome.

|  |  | **OR (95% CI)** | **p** |
| --- | --- | --- | --- |
| **PICU Hospitalization** | No | Reference |  |
|  | Yes | 1.06 (1, 1.13) | 0.067 |
| **Age** | 3 – 5 | 0.64 (0.6, 0.69) | <.001 |
|  | 6 – 10 | 0.89 (0.84, 0.96) | 0.001 |
|  | 11 – 14 | 1.06 (0.99, 1.13) | 0.096 |
|  | 15 – 18 | Reference |  |
| **Sex** | Female | Reference |  |
|  | Male | 0.97 (0.92, 1.01) | 0.161 |
| **Number of CCCs** | 0 | Reference |  |
|  | 1 | 1.17 (1.1, 1.24) | <.001 |
|  | 2 | 1.11 (1.02, 1.22) | 0.021 |
|  | 3+ | 1.15 (1, 1.32) | 0.059 |
| **Hospital Length of Stay, days** | 0 – 2 | Reference |  |
|  | 3 – 7 | 1.03 (0.97, 1.08) | 0.337 |
|  | 8 – 14 | 1.33 (1.2, 1.48) | <.001 |
|  | 15 – 30 | 1.75 (1.5, 2.04) | <.001 |
|  | 31+ | 2.67 (2.13, 3.36) | <.001 |
| **Level of Support** | None | Reference |  |
|  | Mech Vent Only | 0.94 (0.78, 1.12) | 0.469 |
|  | CRRT Only | 0.66 (0.21, 2.13) | 0.492 |
|  | Mech Vent & CRRT | 1.8 (0.21, 15.22) | 0.588 |

Abbreviations: CCC, chronic complex condition; CRRT, continuous renal replacement therapy; PICU, pediatric intensive care unit Model was also adjusted for an additional independent variable Major Diagnosis Category (MDC).

### Secondary Outcomes

Regardless of PICU hospitalization status, patients participated in an average of 2 visits with a new mental health provider over the year following hospitalization. Patients with an index PICU hospitalization were more likely to participate in mental health visits conducted by supportive therapists and psychologists, and less likely to participate in mental health visits conducted by psychiatric prescribers when compared to patients hospitalized on the floor (Table 2). Patients with a PICU hospitalization were less likely to be prescribed new psychiatric medications across nearly all drug classes, including antidepressants, antipsychotics, stimulants, and anxiolytics (Table 4). However, patients with a PICU hospitalization were more likely than floor patients to be prescribed barbiturates, benzodiazepines, and mood-stabilizers.

**Table 4.** Psychopharmacy prescriptions filled within one year following discharge.

| Therapeutic Class <sup>a</sup> | Overall<br>(N=144763) | Non-PICU Hospitalizations<br>(N=114769, 79.3%) |  | PICU Hospitalizations<br>(N=29994, 20.7%) |  |
| --- | --- | --- | --- | --- | --- |
|  |  | No Post-Hospital MH<br>Visits<br>(N=104950, 91.4%) | Post-Hospital MH<br>Visits<br>(N=9819, 8.6%) | No Post-Hospital MH<br>Visits<br>(N=27074, 90.3%) | Post-Hospital MH Visits<br>(N=2920, 9.7%) |
| Stimulant, Amphetamine Type | 13182 (9.1) | 8072 (7.7) | 2527 (25.7) | 1857 (6.9) | 726 (24.9) |
| ASH, Benzodiazepines | 12391 (8.6) | 7798 (7.4) | 1132 (11.5) | 3028 (11.2) | 433 (14.8) |
| Antidepressants | 12313 (8.5) | 7151 (6.8) | 2761 (28.1) | 1639 (6.1) | 762 (26.1) |
| Anxiolytic/Sedative/Hypnotic NEC | 7199 (5) | 4583 (4.4) | 1230 (12.5) | 1035 (3.8) | 351 (12) |
| Psychother, Tranq/Antipsychotics | 5193 (3.6) | 2697 (2.6) | 1420 (14.5) | 665 (2.5) | 411 (14.1) |
| ASH, Barbiturates | 670 (0.5) | 349 (0.3) | 30 (0.3) | 277 (1) | 14 (0.5) |
| Antimanic Agents, NEC | 187 (0.1) | 65 (0.1) | 71 (0.7) | 21 (0.1) | 30 (1) |

All percentages are given as percentage of the column sample except in the header rows, where they are percentage of the larger population.
All features are significantly associated with Post-Hospital MH visits, within each PICU vs Non-PICU Hospitalization ( $p < 0.01$ ), except for ASH, Barbiturates among Non-PICU Hospitalizations.
Abbreviations: ASH, anxiolytic / sedative / hypnotic; PICU, pediatric intensive Care Unit; MH, mental health; NEC, not elsewhere classified
<sup>a</sup> Therapeutic classes were defined from the Merative Marketscan Medicaid Dataset Dictionary and manually selected for psychopharmacy classes. These derived groupings are based on an aggregation of the American Hospital Formulary Service Classification Compilation (AHFSCC) therapeutic class.

The rate of new mental health diagnoses did not differ between those with PICU exposure and without PICU exposure for most diagnostic categories, including internalizing disorders, neurocognitive impairment, neurodevelopmental disorders, severe mental illness, or suicidal behaviors. PICU patients were less likely to be diagnosed with externalizing disorders than patients without a PICU hospitalization (Table 2). However, when adjusted for PICU exposure, sex, number of CCCs, and level of support received, PICU hospitalization was significantly associated with new mental health follow-up for internalizing disorders (OR 1.14 [95% CI: 1.05, 1.24], p=0.002) and for externalizing disorders (OR 1.14 [95% CI: 1.01, 1.28], p=0.03) compared to non-PICU hospitalizations (Supplementary Table 5).

## Discussion

In this study of nearly 150,000 Medicaid-covered pediatric hospitalizations, only 5.2% of patients attended a new mental health care follow-up visit post-discharge. Although a greater proportion of patients with PICU hospitalizations had new mental health visits than non-PICU hospitalizations, the presence of a PICU stay during the hospitalization was not independently associated with new mental health follow-up, though it was associated with new internalizing and externalizing diagnoses. This follow-up rate of 5.2% is lower than reported rates of new mental health symptoms and diagnoses in several population-based cohort studies,^28–33^ suggesting ongoing under-recognition of psychosocial impacts of hospitalization on children and families or other barriers to receiving follow-up care.

Only two other studies have attempted to quantify new mental health care received following pediatric hospitalizations with or without PICU stay.^10^ Daughtrey and colleagues used a commercial payor database and examined the proportion of patients receiving new mental health diagnoses. They observed 6.9% of patients receiving a new mental health diagnosis following hospitalization with a greater proportion (8.3%) from hospitalizations including an ICU stay.

Similar to our findings, they reported higher risk of new mental health diagnoses among children with complex chronic diseases.^34^ Several factors likely contribute to these different observations. Diagnoses are not the same as visits to a mental health provider and may include diagnoses made by primary care pediatricians that would not have been included in our study. There may also be resource differences among children with commercial insurance versus Medicaid insurance which may impact mental health follow-up. Finally, our study excluded patients with known mental health admissions and with TBI admissions, which may influence post-discharge diagnoses and visits. These exclusions may have been balanced by inclusion of some visits providing family-focused mental health care to parents of children. Logan and colleagues reported that 9.5% of parents of PICU survivors received new mental health diagnoses following their child’s admission,^35^ reinforcing the notion that the impact of a pediatric ICU hospitalization impacts not just the survivor but the whole family.^36^

Numerous retrospective and prospective studies have shown that recovery from critical illness is complicated by emotional health sequelae, both for survivors and families.^6,7,37^ However, only pre-hospital trauma has consistently predicted risk of post-intensive care psychological distress.^8,38^ Features of the PICU such as number of invasive procedures, severity of illness, and length of PICU stay have been associated with specific positive or negative emotional health outcomes but studies have been limited by sample size, single-center design, disease specificity, and parental inclusion and findings are inconsistent across studies.^7,39–43^ While we could not measure granular features of the PICU stay in this administrative dataset, we observed no significant associations between level of organ support, including mechanical ventilation and continuous renal replacement therapy. We did find a significant dose-response relationship with hospital length of stay and odds of new mental health visit. This is consistent with a prior study linking longer length of stay with more symptoms of post-traumatic stress disorder in children.^41^ However, two other observational studies have suggested that longer length of stay may be protective and indicative of post-traumatic resiliance,^44,45^ but this may not apply equally to all populations.^43^ Additional research clarifying associations between granular features of the PICU stay and emotional health outcomes are necessary.

Hospitalization that included a PICU stay was significantly associated with new diagnoses of both internalizing and externalizing disorders. Post-traumatic stress in children can present as a wide spectrum of emotional and behavioral difficulties, including internalizing symptoms such as anxiety, depression, withdrawal, or regression in developmental milestones as well as externalizing symptoms like aggression, irritability, or oppositional or risky behaviors.^46^ These may be coded separately or as part of the syndrome of post-traumatic stress, such that the diagnostic variability we observed may reflect the diverse clinical expression of trauma-related distress in this population.

Notably, to treat these new diagnoses, PICU patients were more likely than floor patients to see talk therapists (i.e., psychologists or supportive therapists) whereas floor patients were more likely to see psychiatric prescribers. It is possible that these observed differences in provider visits are primarily driven by systemic, structural, and personal barriers. Despite well documented links between hospitalization and increased emotional distress,^6^ standards for screening and referral to mental health providers are lacking.^21^ Moreover, medical and mental health care are often siloed, preventing effective care coordination.^47^ General guidelines for referral to pediatric psychiatrists versus talk therapists are emerging; most suggest talk therapy as a first line treatment for mild to moderate emotional symptoms that are not complicated by high-risk behaviors (e.g., suicidality).^48,49^ However, in practice, referral sources are often constrained by structural and personal barriers. Structural barriers, such as lack of access to affordable, high quality care and a severe shortage of mental health care providers, may discourage providers from making referrals and/or prevent families from connecting with services.^50,51^ Embedding mental health providers in primary care or follow-up clinics has been shown to facilitate care coordination and improve access to care.^52,53^ This may particularly improve access in children recovering from critical illness, when families may have a large number of follow up appointments and therefore deprioritize mental health visits.^52^Critically, given potential changes to Medicaid funding availability, accessing mental health care may become increasingly challenging, even beyond what was documented in the current findings. Numerous familial and patient characteristics may also interfere with effective referral, including mental health stigma, cultural beliefs around mental health care, lack of trust in mental health systems, and inadequate emotional or psychological readiness to accept mental health care.^54–58^ Parental factors, including parental mental health sequelae related to their children’s hospitalization or parents de-prioritizing mental health visits due to overwhelming medical needs may also interfere with access to care.^35^ Notably, even in under-resourced settings, adherence rates for medical follow-up after pediatric intensive care is generally high (>65%),^59–61^ indicating that effective referral to mental health care is possible if these barriers can be identified and addressed.^21^ Future studies are needed to explicitly examine, identify, and address barriers to effective screening and referral, for example by considering access to telehealth appointments, embedded mental health providers in clinic, or through digital health focused interventions.^62,63^

That PICU patients were more likely that floor patients to be prescribed barbiturates and benzodiazepines may reflect their use as medical sedatives, to treat seizure, or to complete habituation weans post-discharge rather than their use as primary anxiolytics; however, we could not test that hypothesis in our study.^64,65^ Future studies are needed to examine how and why psychiatric medications are prescribed in PICU patients.

Finally, it is important to note that PICU hospitalization did not emerge as a unique predictor of new mental health follow up among the specific patients included in this study. This finding may reflect the resilience of children in that the majority of children exposed to life-threatening trauma do not go on to develop posttraumatic stress disorder or other significant mental health concerns.^66^ Individual vulnerability, developmental stage, family context, and post-discharge support all influence mental health symptoms post hospitalization.^6,66^

### Limitations

Our results carry several limitations. While this study used multi-state Medicaid administrative data, we were limited to visits and prescription claims billed through Medicaid and could not identify additional care paid for out-of-pocket or by private insurance, potentially leading to under-estimated care. Similarly, we could not primarily assess the quality of the Medicaid dataset or report details such as missing visits or loss to follow-up. We excluded admissions with primary diagnosis codes related to mental illness; however, we did not exclude for other codes that might plausibly related to mental health (e.g., accidental poisoning). As such, our observed rate for follow-up mental health care may be artificially inflated by patients who were hospitalized for suicidal or non-suicidal self-injurious behaviors.

## Conclusions

Given well-documented associations between medical hospitalization and adverse emotional health outcomes, strikingly few patients completed visits to any mental health care provider in the year following their hospital discharge. Multiple systemic, structural, and personal barriers likely contribute to this gap in care. Establishing consistent guidelines, implementing routine screening, and making proactive referrals to community providers represent critical first steps toward improving post-hospital mental health linkage for children and families.

## Author Contributions

Drs. Mariah DeSerisy, Julia Heneghan, and Adam Dziorny conceptualized the study, developed the analysis plan, interpreted results, drafted the manuscript, and critically reviewed and revised the manuscript.

Dr. Matt Hall developed the analysis plan, accessed the data, conducted the analysis, interpreted results, and critically reviewed and revised the manuscript.

Drs. Daniel Choi, Leslie Dervan, Daniel Garros, Denise Goodman, Jason Kane, Joseph Kohne, Colin Rogerson, Nadia Roumeliotis, and Vanessa Toomey developed the analysis plan, interpreted results, and critically reviewed and revised the manuscript.

All authors approved the final manuscript as submitted and agree to be accountable for all aspects of the work including the data, analysis, and conclusions.

## Conflict of interest disclosure and source of funding

The authors have no conflicts of interest relevant to this article to disclose. No dedicated source of funding supported this work.

## Data Availability

All data produced in the present study are available upon reasonable request to the authors.

## Notes

### Competing Interest Statement

The authors have declared no competing interest.

### Funding Statement

This study did not receive any funding.

### Author Declarations

IRB of the University of Rochester waived ethical approval for this work (Study # 00010686) and determined that the proposed activity is not research involving human subjects as defined by DHHS and FDA regulations.

