## Supplemental Data for "Prevalence and Characteristics of New Mental Health Interventions in PICU Survivors"

**Supplementary Figure 1.** Patient flow diagram

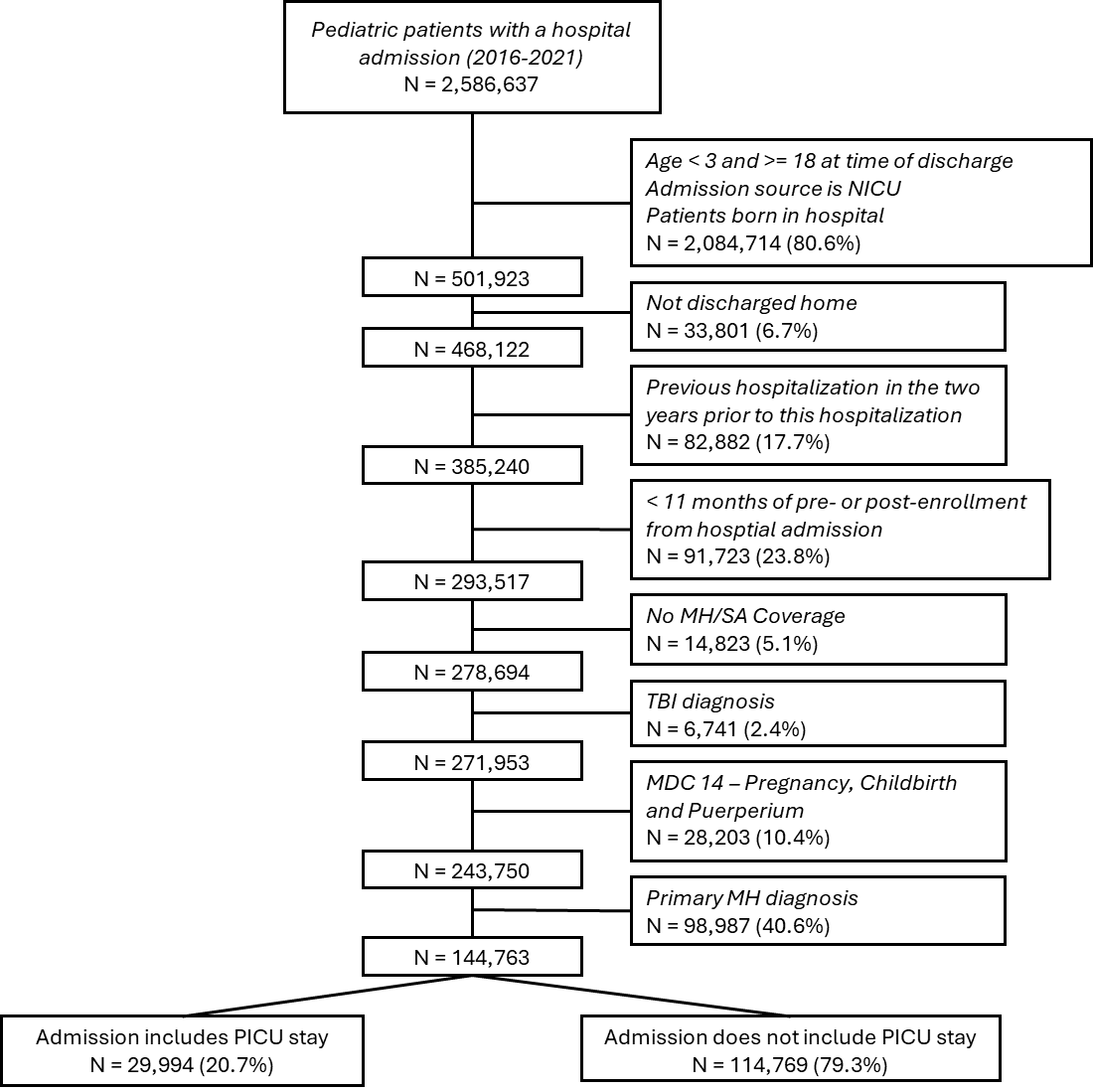

Abbreviations: MDC, major diagnostic category; MH, mental health; NICU, neonatal intensive care unit; SA, substance abuse; TBI, traumatic brain injury

**Supplementary Table 2.** International Classification of Diseases (ICD) codes used to identify mechanical ventilation and / or continuous dialysis.

| **Mechanical Ventilation** | **Continuous Dialysis** |
| --- | --- |
| ICD-9-CM code 96.7x  ICD-10-CM codes 5A1935Z, 5A1945Z, and 5A1955Z | ICD-9 code 39.95  ICD-10 codes 5A1D00Z, and 5A1D60Z |

**Supplementary Table 3.** Top 20 conditions (PECCS) by ICU admission status. Shaded rows are those diagnoses with >10% post-hospitalization MH utilization.

| **No PICU Admission During Hospitalization** | | |  | **PICU Admission During Hospitalization** | | |
| --- | --- | --- | --- | --- | --- | --- |
| **Category Description** | **N** | **% with Post-Hospitalization Mental Health Utilization** |  | **Category Description** | **N** | **% with Post-Hospitalization Mental Health Utilization** |
| Asthma | 10935 | 4.9 |  | Asthma | 4034 | 6.0 |
| Pneumonia | 6973 | 5.9 |  | Diabetic ketoacidosis | 2893 | 10.9 |
| Acute appendicitis with peritonitis | 5368 | 5.4 |  | Respiratory failure; insufficiency; arrest | 2052 | 6.6 |
| Cellulitis | 5189 | 7.2 |  | Seizures w and w/o intractable epilepsy | 1201 | 13.3 |
| Urinary tract infections | 3227 | 8.2 |  | Pneumonia | 1129 | 5.1 |
| Dehydration | 2598 | 8.4 |  | Scoliosis | 968 | 7.9 |
| Seizures w and w/o intractable epilepsy | 2487 | 14.5 |  | Septicemia (except in labor) | 853 | 8.8 |
| Fracture of lower limb | 2480 | 6.5 |  | Crushing injury or internal injury | 696 | 11.9 |
| Constipation | 2183 | 13.7 |  | Burns | 667 | 8.2 |
| Acute appendicitis w/o peritonitis | 1956 | 6.7 |  | Partial epilepsy w and w/o intractable epilepsy | 479 | 13.6 |
| Sickle cell disease with crisis | 1856 | 8.9 |  | Poisoning by other medications and drugs | 461 | 16.5 |
| Diabetic ketoacidosis | 1849 | 9.4 |  | Complication of device; implant or graft | 342 | 11.1 |
| Gastroenteritis, infectious | 1710 | 7.9 |  | Compression of brain | 320 | 13.4 |
| Scoliosis | 1652 | 8.3 |  | Other convulsions | 281 | 18.1 |
| Partial epilepsy w and w/o intractable epilepsy | 1371 | 15.7 |  | Acute bronchiolitis | 273 | 5.1 |
| Viral infection | 1318 | 7.0 |  | Diabetes mellitus with complications | 265 | 10.2 |
| Burns | 1254 | 7.7 |  | Viral Infection (COVID-19) | 254 | 5.9 |
| Septicemia (except in labor) | 1191 | 8.0 |  | Epilepsy; convulsions | 249 | 11.6 |
| Impaction of intestine | 1121 | 16.4 |  | Ostium secundum atrial septal defect | 234 | 6.8 |
| Type 1 diabetes mellitus with complications | 1109 | 13.2 |  | Other nervous system disorders | 214 | 16.8 |

Abbreviations: MH, mental health; PICU, pediatric intensive care unit

**Supplementary Table 4**. Mental health provider utilization within one year following index admission, including patients with MH utilization pre-hospitalization.

|  |  | **Overall**  **(N=12739)** | **Non-PICU Hospitalization**  **(N=9819)** | **PICU Hospitalization**  **(N=2920)** | **p-value** |
| --- | --- | --- | --- | --- | --- |
| **Number of visits to MH provider** | Median [IQR] | 4 [1, 10] | 4 [1, 10] | 3 [1, 9] | 0.011 |
| **Rate of MH visit to MH provider, per 100 children** |  | 78.9 (78.45, 79.36) | 78.4 (77.9, 78.9) | 80.9 (79.9, 82) | <.001 |
| **Rate of MH visit by provider type, per 100 children** | Psychiatry | 17.91 (17.69, 18.13) | 18.6 (18.4, 18.9) | 15.2 (14.7, 15.6) | <.001 |
|  | Therapists (Supportive) | 43.74 (43.4, 44.08) | 42.9 (42.5, 43.3) | 47 (46.2, 47.8) | <.001 |
|  | Psychologist | 17.25 (17.04, 17.47) | 16.8 (16.6, 17.1) | 18.8 (18.3, 19.3) | <.001 |
| **MH Visit Diagnosis** | Internalizing | 6779 (53.2) | 5219 (53.2) | 1560 (53.4) | 0.796 |
|  | Externalizing | 4786 (37.6) | 3762 (38.3) | 1024 (35.1) | 0.002 |
|  | Neurodevelopmental Disorders | 2283 (17.9) | 1752 (17.8) | 531 (18.2) | 0.672 |
|  | Neurocognitive Impairment | 802 (6.3) | 620 (6.3) | 182 (6.2) | 0.874 |
|  | Severe Mental Illness | 505 (4) | 386 (3.9) | 119 (4.1) | 0.726 |
|  | Suicidal Behaviors | 71 (0.6) | 58 (0.6) | 13 (0.4) | 0.354 |
|  | Other | 748 (5.9) | 584 (5.9) | 164 (5.6) | 0.504 |

Abbreviations: ICU, Intensive Care Unit; MH, mental health

**Supplementary Table 5**. Multivariable logistic regression model of secondary outcomes.

|  |  | **MH follow-up with the diagnosis of an Internalizing disorder** | | **MH follow-up with the diagnosis of an Externalizing disorder** | |
| --- | --- | --- | --- | --- | --- |
|  |  | **OR (95% CI)** | **p** | **OR (95% CI)** | **p** |
| **ICU Utilization** | No | Reference | | | |
|  | Yes | 1.14 (1.05, 1.24) | 0.002 | 1.14 (1.01, 1.28) | 0.030 |
| **Age** | 3 – 5 | 0.3 (0.27, 0.34) | <.001 | 0.61 (0.52, 0.7) | <.001 |
|  | 6 – 10 | 0.62 (0.56, 0.67) | <.001 | 1.4 (1.24, 1.58) | <.001 |
|  | 11 – 14 | 0.95 (0.87, 1.04) | 0.254 | 1.18 (1.03, 1.35) | 0.015 |
|  | 15 – 18 | Reference | | | |
| **Sex** | Female | Reference | | | |
|  | Male | 0.66 (0.62, 0.71) | <.001 | 1.55 (1.41, 1.7) | <.001 |
| **Number of CCCs** | 0 | Reference | | | |
|  | 1 | 1.04 (0.96, 1.12) | 0.381 | 0.95 (0.85, 1.06) | 0.380 |
|  | 2 | 0.84 (0.73, 0.95) | 0.008 | 0.65 (0.53, 0.8) | <.001 |
|  | 3+ | 0.69 (0.54, 0.87) | 0.002 | 0.45 (0.3, 0.68) | <.001 |
| **Hospital Length of Stay, days** | 0 – 2 | Reference | | | |
|  | 3 – 7 | 1.01 (0.94, 1.09) | 0.753 | 0.97 (0.88, 1.07) | 0.491 |
|  | 8 – 14 | 1.53 (1.33, 1.75) | <.001 | 0.97 (0.77, 1.22) | 0.794 |
|  | 15 – 30 | 1.81 (1.47, 2.24) | <.001 | 0.8 (0.52, 1.22) | 0.303 |
|  | 31+ | 2.96 (2.2, 3.99) | <.001 | 1.74 (1.01, 3.02) | 0.046 |
| **Level of Support** | None | Reference | | | |
|  | Mech Vent Only | 0.83 (0.63, 1.08) | 0.171 | 0.74 (0.49, 1.12) | 0.153 |
|  | CRRT Only | 0.87 (0.21, 3.59) | 0.849 | - ^a^ |  |
|  | Mech Vent & CRRT | 3.15 (0.37, 27.13) | 0.297 | - ^a^ |  |

Abbreviations: CCC, chronic complex condition; CRRT, continuous renal replacement therapy; ICU, intensive care unit

Model was also adjusted for an additional independent variable Major Diagnosis Category (MDC).

^a^ Insufficient number of patients to include these categories.
